# Oral Hygiene Practices and Their Impact on Oral Health Among the Indigenes of Akpugo Community in Enugu State, Nigeria

**DOI:** 10.1101/2024.06.21.24309281

**Authors:** Chisom Promise Nnamani, Muneer Yaqub

**Author notes:** **Corresponding Author:** Muneer Yaqub. Address: 800 W Campbell Rd, Richardson, TX 75080. **Disclosure Statement:** There are no financial conflicts of interest to disclose.

## Abstract

**Objective:** This study aims to evaluate the oral hygiene practices and their effectiveness in preventing common oral diseases among the residents of the Akpugo community in Enugu State, Nigeria.

**Methods:** A cross-sectional survey was conducted among 150 randomly selected participants from the five villages of Akpugo. Data collection involved personal interviews and physical examinations of oral cavities using sterilized instruments. The analysis included percentage calculations, z-tests, and the categorization of participants by socio-economic groups based on age and gender.

**Results:** The study revealed a high prevalence of oral diseases, with 29.3% of participants having dental caries, 19.3% suffering from halitosis, 18% diagnosed with gingivitis, and 10% with periodontitis. Most participants (90.7%) brushed their teeth once daily, predominantly in the morning, and only 5.3% brushed twice daily. A majority used toothbrushes and toothpaste (58%), while a significant portion still relied on traditional methods such as chewing sticks (26.67%). The presence of dental calculus was observed in 47.3% of participants, with dental plaque noted in 16.7%.

**Conclusion:** The findings highlight a significant prevalence of poor oral health despite varying levels of awareness regarding preventive practices. The study underscores the need for targeted interventions, including comprehensive oral health education programs, the establishment of dental centers, and community-led initiatives to improve oral hygiene practices. These measures are crucial for reducing the incidence of oral diseases and enhancing the overall health and well-being of the Akpugo community. The study emphasizes the importance of continuous education and intervention to promote optimal oral health in rural areas.

## INTRODUCTION

In contemporary health discourse, the prevalence of various diseases that impact humans is a significant concern, particularly those caused by pathogenic microorganisms. These infections can affect any part of the body, and their spread is often facilitated by inadequate personal hygiene practices. One critical area highly susceptible to such infections is the oral cavity, which is constantly exposed to a myriad of external factors, making it prone to various oral diseases [1].

The oral cavity, located within the head region, is not only essential for maintaining facial aesthetics but also plays a crucial role in fundamental activities such as mastication, speech, and respiration. However, its continuous exposure to diverse external elements, particularly through the intake of food and other substances, makes it a primary target for pathogenic microorganisms. These organisms interact with dental plaque and food debris on the teeth surfaces, metabolizing sugars and starches to produce acids. These acids can demineralize the tooth enamel and damage the supporting structures, leading to common oral diseases such as dental caries (tooth decay), gingivitis (gum inflammation), periodontitis (advanced gum disease), and halitosis (bad breath) [2]. The prevention of these conditions hinges on the implementation of effective oral hygiene practices.

Oral hygiene encompasses the systematic practice of maintaining the cleanliness of the oral cavity to prevent diseases and other problems such as bad breath. This involves regular tooth brushing and the adoption of other good hygiene habits. The principal objective of these practices is to create an oral environment that is unfavorable for pathogenic microorganisms, thereby preventing the development of oral diseases and promoting long-term dental health [3].

Despite advances in education and increased health awareness, many individuals, especially those in rural communities, do not engage in basic oral hygiene practices. Numerous studies have indicated a general lack of awareness regarding oral health, irregular tooth brushing habits, and overall poor oral hygiene among populations in Nigeria [4]. This negligence often leads to a high incidence of oral diseases, with affected individuals frequently resorting to traditional remedies and self-medication. This trend is particularly noticeable in rural areas such as the Akpugo community in Enugu State [5].

This study aims to evaluate the oral hygiene practices among the residents of the Akpugo community and their effectiveness in preventing oral diseases. By conducting a thorough assessment and providing targeted educational interventions, this research seeks to promote improved oral health practices within the community. The study employs a cross-sectional survey design to collect data on current oral hygiene habits and the prevalence of oral diseases among the residents. The methodology includes personal interviews, physical examinations of the oral cavities using sterilized instruments, and rigorous validation of the data collection tools. The data are analyzed using statistical methods and presented in tabular form to illustrate the distribution of various oral hygiene practices and the prevalence of oral diseases. Additionally, the study incorporates educational interventions, such as oral hygiene education and tooth brushing demonstrations, to enhance the participants’ knowledge and practice of good oral hygiene.

The findings from this study are expected to provide critical insights into the oral health status of the Akpugo community. By identifying gaps in current practices and recommending effective measures, the study aims to improve oral health awareness and hygiene practices. The establishment of dental centers and the implementation of targeted oral health campaigns and seminars are recommended to foster a culture of good oral hygiene within the community. These initiatives are vital for reducing the prevalence of oral diseases and enhancing the overall health and well-being of the Akpugo residents. In conclusion, this research underscores the importance of adopting effective oral hygiene practices to prevent common oral diseases. It highlights the necessity for continuous education and intervention to promote optimal oral health in rural communities like Akpugo. Through comprehensive assessment and targeted educational initiatives, this study aims to empower the community with the knowledge and tools required to maintain good oral health and improve their quality of life.

## MATERIAL AND METHODS

### Study Design and Setting

A cross-sectional survey design was employed to assess oral hygiene practices among the indigenes of the Akpugo community in Enugu State, Nigeria. Akpugo, located within the Nkanu West Local Government Area, comprises a population of approximately 4000 individuals across five villages, predominantly inhabited by civil servants, students, traders, and businesspeople [6]. Population distribution and demographics were sourced from His Royal Highness Igwe Michael Chukwudike.

### Sampling and Participants

A sample of 150 inhabitants was selected using a simple random sampling technique, with 30 individuals chosen from each of the five villages. This approach ensured a representative subset of the community. Inclusion criteria included residents aged 18 and above who consented to participate in the study.

### Data Collection

Data collection involved personal interviews and physical examinations of oral cavities. The instruments used for examinations included mouth mirrors, college tweezers, and caries probes, all sterilized using appropriate cold sterilization chemicals to ensure hygienic procedures.

Interviews: Structured interviews were conducted to gather information on participants’ oral health practices and personal data. Interviews covered topics such as frequency and method of tooth brushing, use of oral hygiene aids, dietary habits, and access to dental care [7].

Oral Examinations: Intra-oral examinations were performed to assess the condition of participants’ oral cavities. Findings, including the presence of dental plaque, calculus, material alba, and dental stains, were recorded on data sheets. Examinations also identified common oral diseases such as dental caries, halitosis, gingivitis, and periodontitis [8].

### Ethical Considerations

Informed consent was obtained from the community leader and all participants prior to data collection. Participants were assured of confidentiality and anonymity. The study was approved by the ethical review board of Usmanu Danfodiyo University [9].

### Data Analysis

The collected data were organized by sex and age. Statistical analysis was performed using a Z-test to compare the prevalence of oral health conditions and practices across different demographic groups. Results were presented in tables, charts, and descriptive essays to facilitate clear understanding and interpretation [10].

### Interventions

As part of the study, oral hygiene education sessions were conducted. These sessions included demonstrations of proper tooth brushing techniques and the use of oral hygiene aids. Educational materials were distributed to reinforce the importance of good oral hygiene practices [11].

### Reliability and Validity

To ensure the reliability and validity of the data collection tools, the instruments were pre-tested in a pilot study involving a small subset of the community. Feedback from the pilot study was used to refine the interview questions and examination procedures [12]. This comprehensive methodological approach ensured the collection of robust and reliable data, providing valuable insights into the oral hygiene practices and oral health status of the Akpugo community.

## RESULTS

### Gender Distribution Reveals Slight Female Majority

The study included 150 participants from five villages in the Akpugo community, with 72 males (48%) and 78 females (52%). Specifically, 30 participants were selected from each village. Gender distribution showed a higher selection of females from Ndi-Uno (18.1%), Ndi-Agu (21.8%), and Orji-Agu (23.1%), while more males were selected from Orji-Uno (23.6%) and Obuno (18.25%). This distribution is detailed in Table 1 and confirmed by Figure 1.

**Figure 1:**
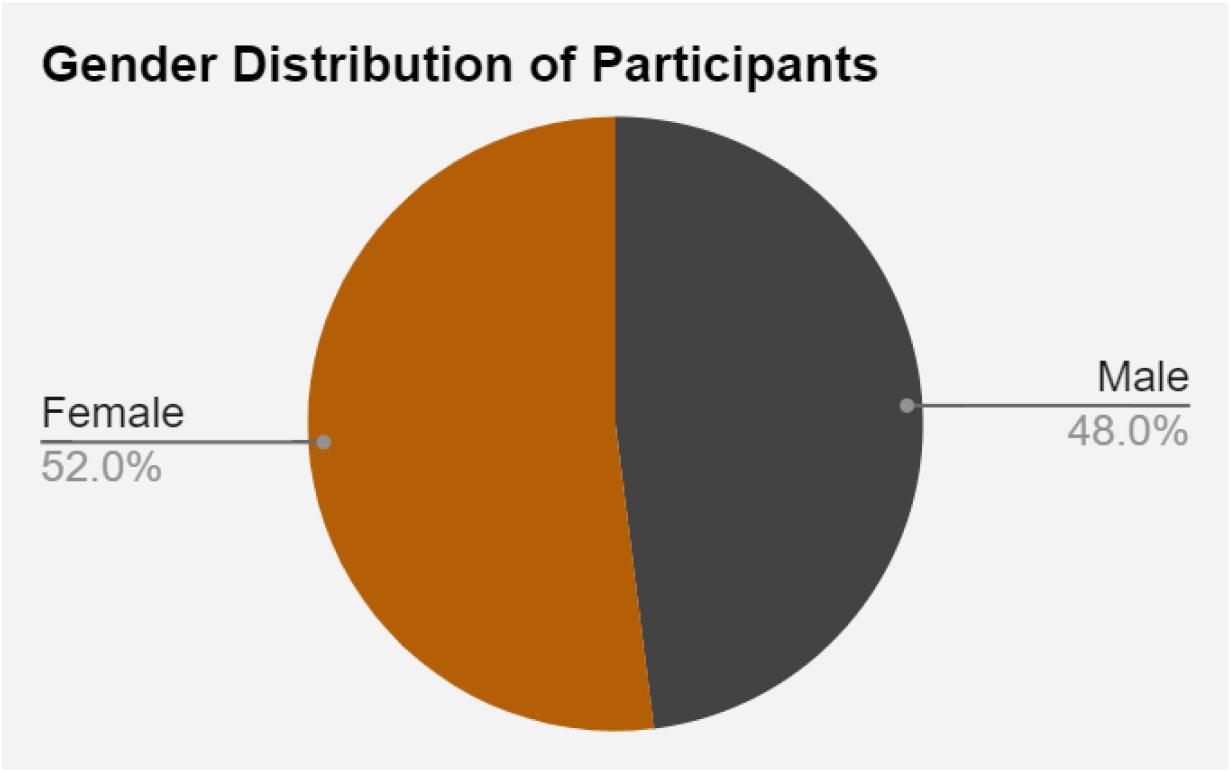
Gender Distribution of the Participants in the Akpugo Community. This figure shows the percentage distribution of male (48%) and female (52%) participants among the 150 individuals sampled from the five villages in the Akpugo community.

**Table 1:** Gender Distribution of Participants. This table details the number and percentage of male and female participants selected from each of the five villages in the Akpugo community.

| **Villages** | **Frequency** | **%** | **No. of**<br>**Males** | **% of**<br>**Males** | **No. of**<br>**Females** | **% of**<br>**Females** |
| --- | --- | --- | --- | --- | --- | --- |
| Ndi-Uno | 30 | 20 | 12 | 16.7 | 18 | 23.1 |
| Ndi-Agu | 30 | 20 | 13 | 18.1 | 17 | 21.8 |
| Orji-Uno | 30 | 20 | 17 | 23.6 | 13 | 16.7 |
| Orji-Agu | 30 | 20 | 12 | 16.7 | 18 | 23.1 |
| Obuno | 30 | 20 | 18 | 25 | 12 | 15.4 |
| **Total** | **150** | **100** | **72** | **100** | **78** | **100** |

**Table 2:** Overall Gender Distribution. This table summarizes the overall gender distribution of the study participants, showing that 48% were males and 52% were females.

| Sex | Frequency | (%) |
| --- | --- | --- |
| Male | 72 | 48 |
| Female | 78 | 52 |
| Total | 150 | 100 |

### Age Group 11-20 Years Has Highest Participant Frequency

The age distribution of participants is depicted in Figure 2 and Table 3. Participants aged 11-20 years had the highest frequency (19.3%), while those aged 71 years and above had the lowest frequency (8%). The detailed distribution was as follows: 13.3% aged 1-10 years, 19.3% aged 11-20 years, 12.7% aged 21-30 years, 13.3% aged 31-40 years, 12.7% aged 41-50 years, 12% aged 51-60 years, 8.7% aged 61-70 years, and 8% aged 71 years and above.

**Figure 2:**
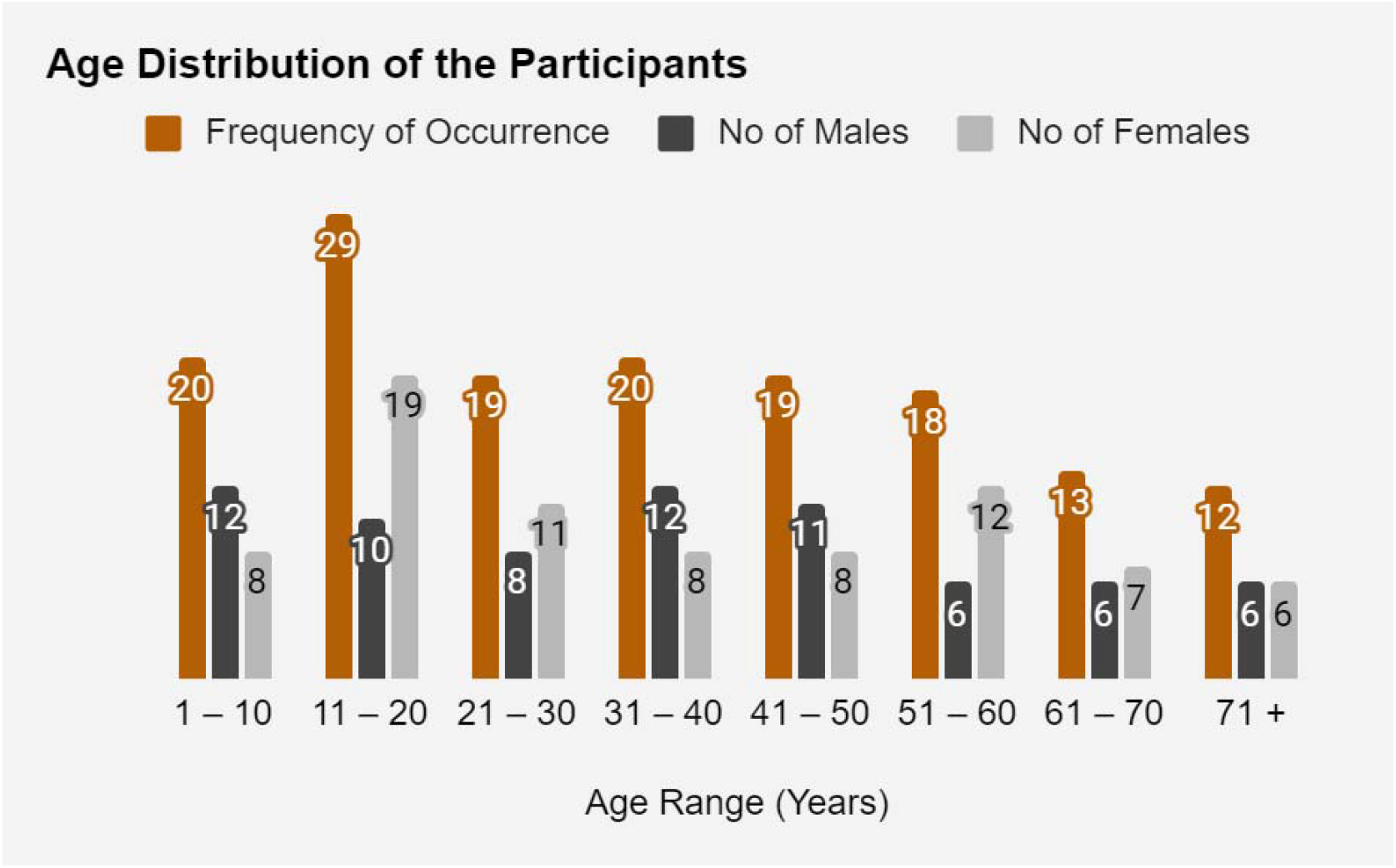
Age Distribution of the Participants. This figure illustrates the age distribution of the participants, highlighting that the highest frequency of participants falls within the 11-20 year age group (19.3%) and the lowest frequency in the 71+ years age group (8%).

**Table 3:**
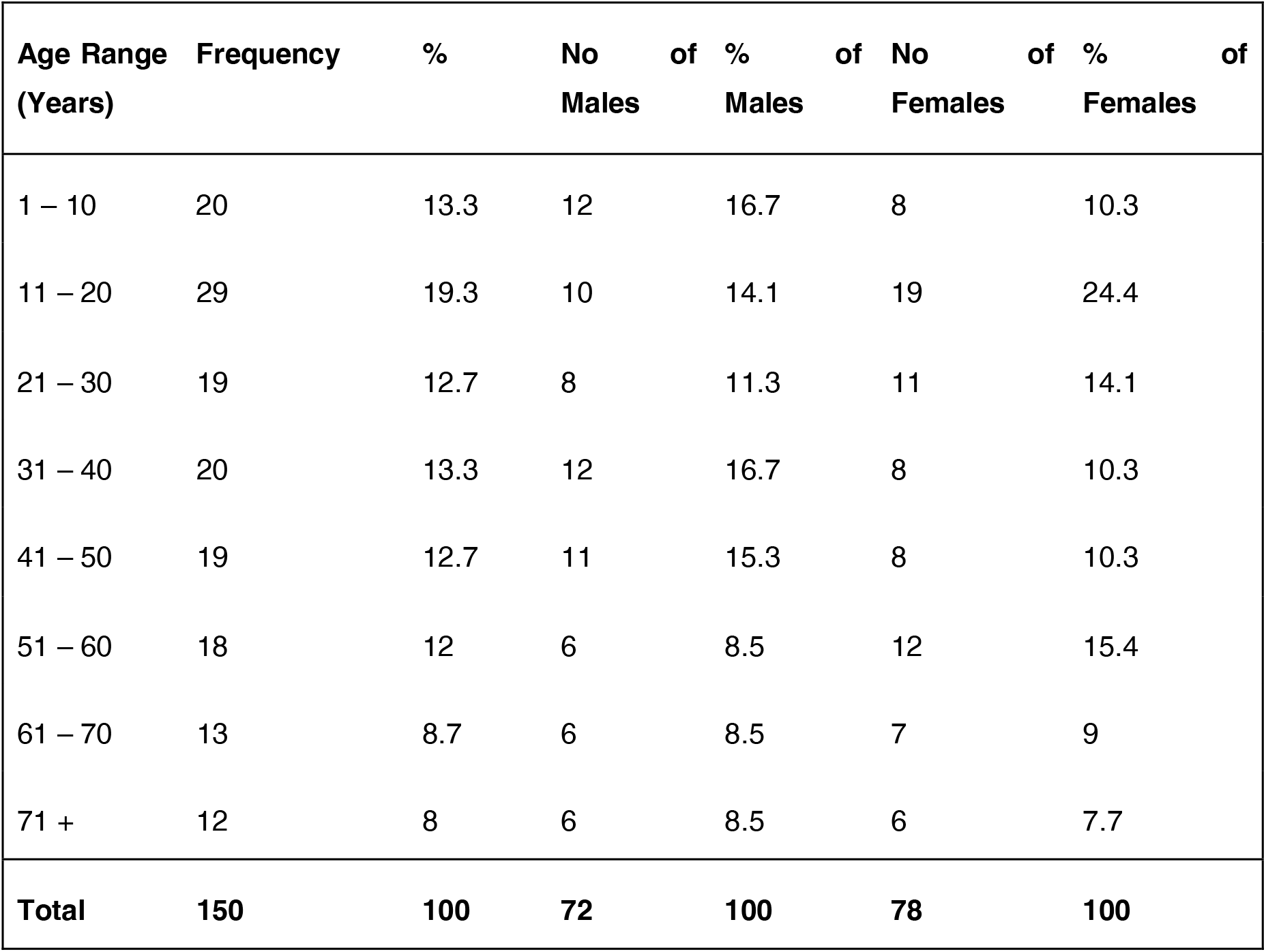
Age Distribution of the Participants. This table provides the number and percentage of participants in each age group, indicating that the 11-20 years age group had the highest representation.

| **Age Range**<br>**(Years)** | **Frequency** | **%** | **No**<br>**of**<br>**Males** | **%**<br>**of**<br>**Males** | **No**<br>**of**<br>**Females** | **%**<br>**of**<br>**Females** |
| --- | --- | --- | --- | --- | --- | --- |
| 1 – 10 | 20 | 13.3 | 12 | 16.7 | 8 | 10.3 |
| 11 – 20 | 29 | 19.3 | 10 | 14.1 | 19 | 24.4 |
| 21 – 30 | 19 | 12.7 | 8 | 11.3 | 11 | 14.1 |
| 31 – 40 | 20 | 13.3 | 12 | 16.7 | 8 | 10.3 |
| 41 – 50 | 19 | 12.7 | 11 | 15.3 | 8 | 10.3 |
| 51 – 60 | 18 | 12 | 6 | 8.5 | 12 | 15.4 |
| 61 – 70 | 13 | 8.7 | 6 | 8.5 | 7 | 9 |
| 71 + | 12 | 8 | 6 | 8.5 | 6 | 7.7 |
| **Total** | **150** | **100** | **72** | **100** | **78** | **100** |

### Majority of Participants Are Farmers

Occupational data, as shown in Figure 3 and Table 4, indicate that of the 150 respondents, 40% were farmers, 13.3% were civil servants, 20% were either businessmen or women, and 26.7% were students.

**Figure 3:**
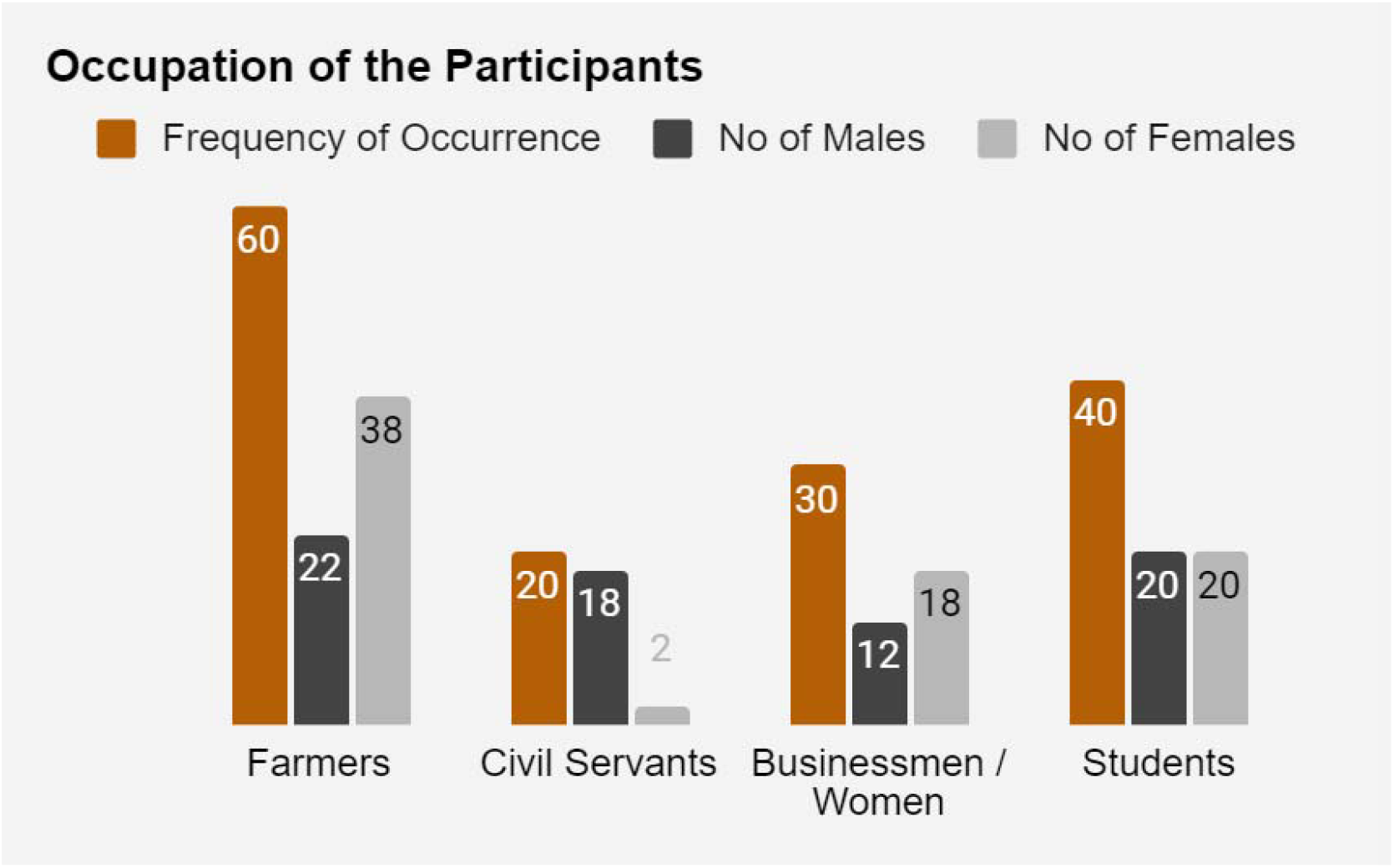
Occupational Distribution of the Participants. This figure depicts the occupational status of the participants, with the majority being farmers (40%), followed by students (26.7%), businessmen/women (20%), and civil servants (13.3%).

**Table 4:**
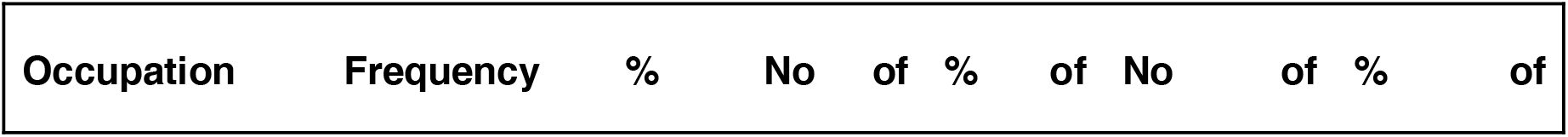
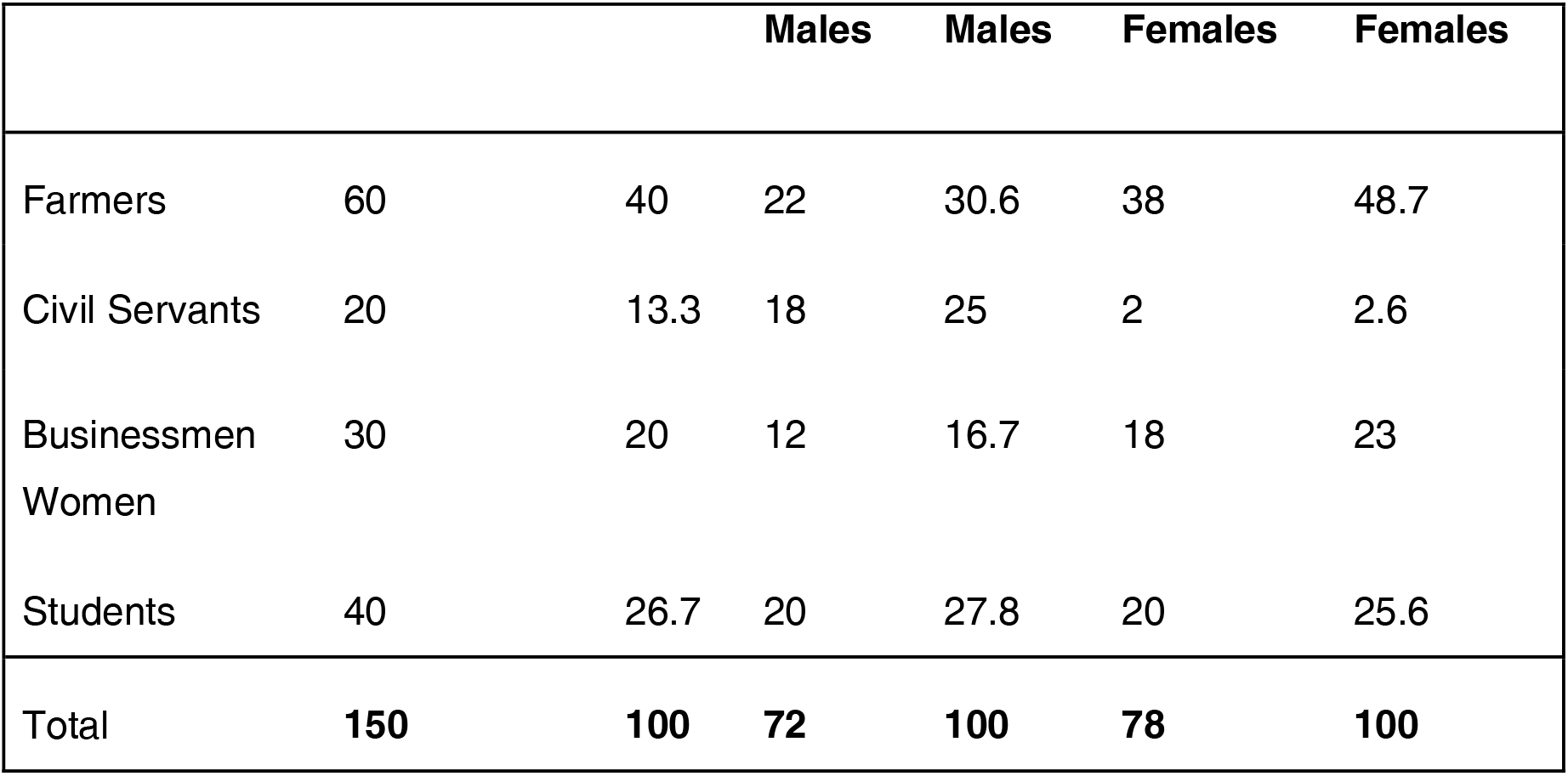
Occupation of the Participants. This table categorizes participants by their occupations, revealing that farming was the predominant occupation among the participants.

### Toothbrush and Paste Are Most Common Oral Hygiene Tools

Oral hygiene measures used by participants across different age groups are illustrated in Figure 4 and Table 5. In the 1-10 years age group, 80% used toothbrush and paste, 10% used chewing sticks, and 10% used their finger and water. For the 11-20 years age group, 85% used toothbrush and paste, and 15% used chewing sticks. Among those aged 21-30 years, 80% used toothbrush and paste, 16.7% used chewing sticks, and 3.3% used water and salt. Participants aged 31-40 years demonstrated varied practices: 47.8% used toothbrush and paste, 21.7% used chewing sticks, 8% used salt and water, and 26.1% used finger and salt. In the age group 41 years and above, 23.5% used toothbrush and paste, 58.8% used chewing sticks, 5.9% used water and salt, and 2.7% used finger and water.

**Figure 4:**
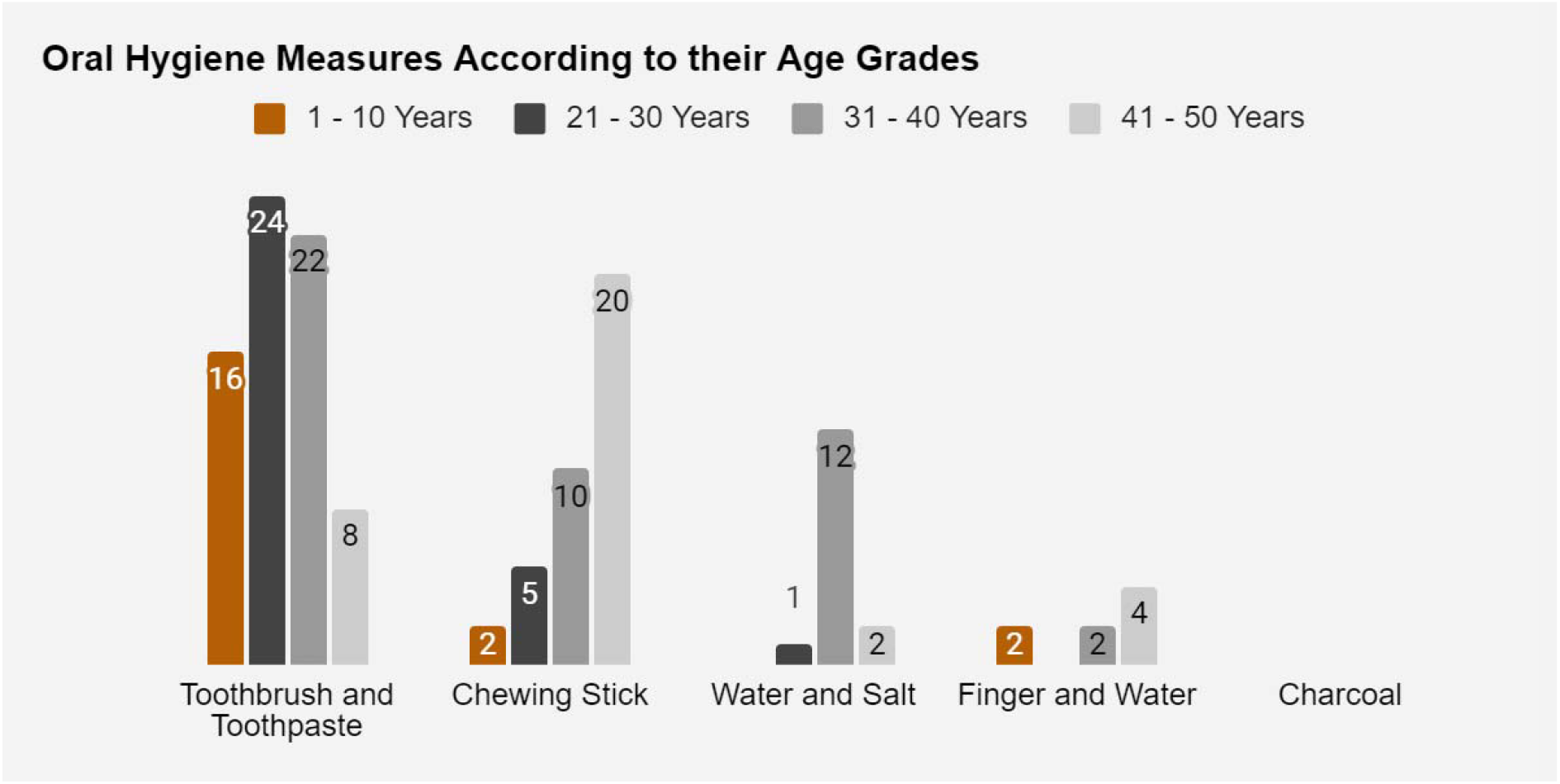
Oral Hygiene Measures by Age Group. This figure shows the various oral hygiene measures used by different age groups, with toothbrush and paste being the most common across all age groups, especially in the 21-30 years age group (80%).

**Table 5:**
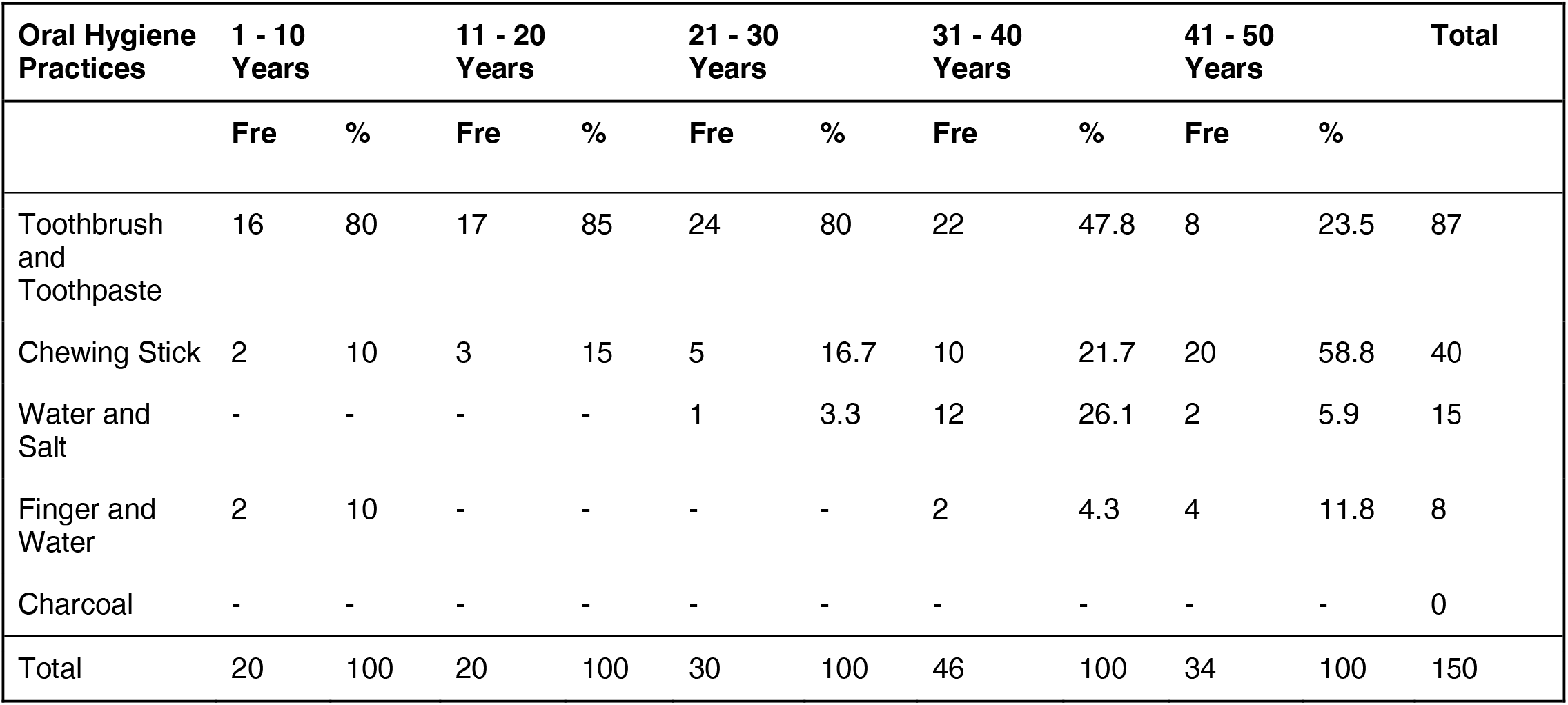
Oral Hygiene Measures Used by Different Age Groups. This table outlines the different oral hygiene measures employed by participants across various age groups, with toothbrush and paste being the most commonly used.

### Morning Brushing Is Most Common Practice

The periods when participants brushed their teeth are presented in Figure 5 and Table 6. A total of 56% brushed in the morning only, with 44.2% being males and 57.7% females. Only 11.3% brushed at night, including 12.5% males and 10.3% females. Additionally, 32.7% brushed both morning and night, comprising 33.3% males and 32.1% females.

**Figure 5:**
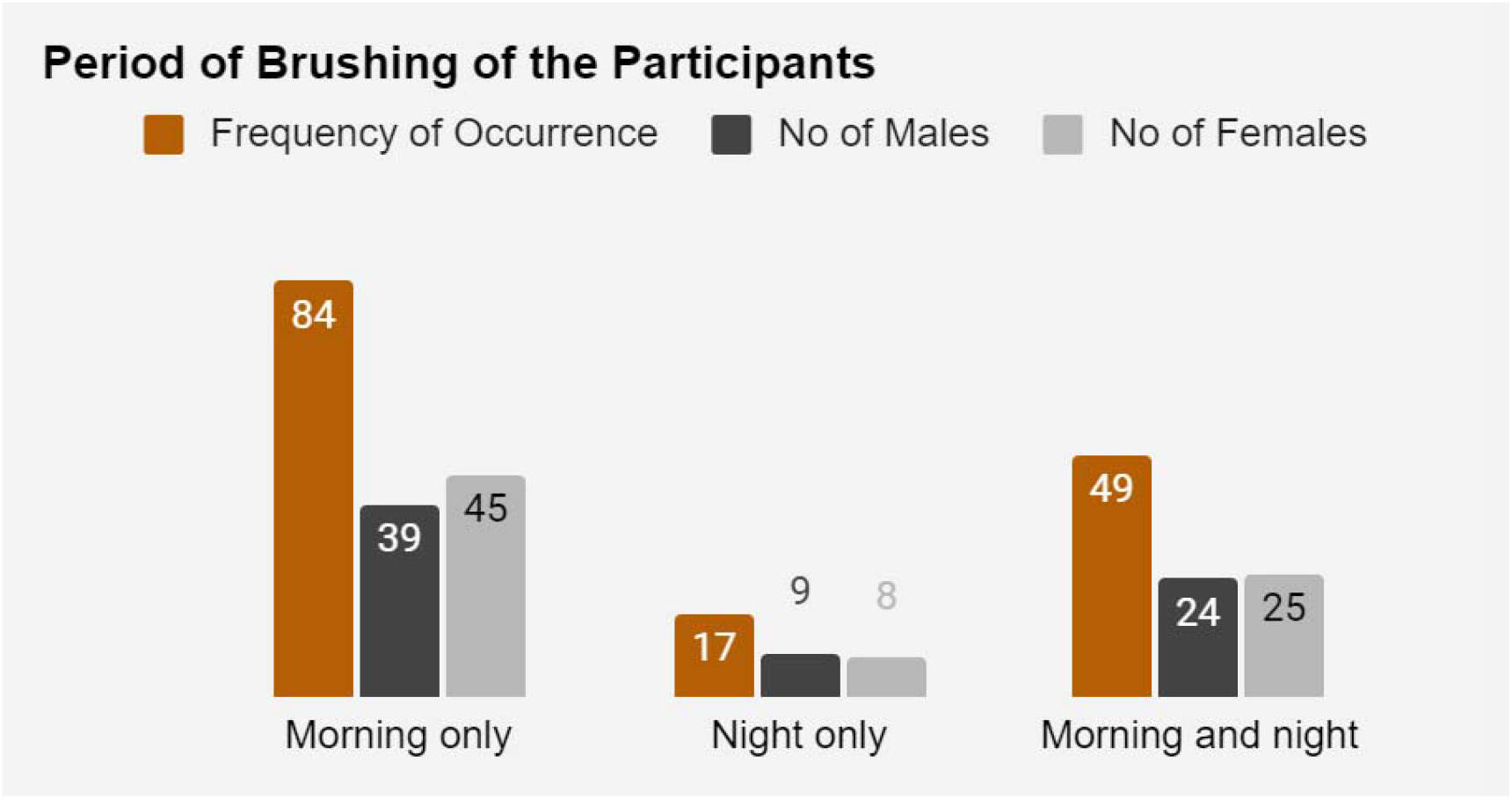
Period of Brushing Among the Participants. This figure displays the times when participants brush their teeth, with 56% brushing in the morning only, 11.3% at night only, and 32.7% both morning and night.

**Table 6:** Period of Brushing. This table shows when participants typically brush their teeth, with the majority brushing only in the morning.

| Period of brushing | Frequency | % | No of Males | % of Males | No of Females | % of Females |
| --- | --- | --- | --- | --- | --- | --- |
| Morning only | 84 | 56 | 39 | 54.2 | 45 | 57.7 |
| Night only | 17 | 11.3 | 9 | 12.5 | 8 | 10.3 |
| Morning and night | 49 | 32.7 | 24 | 33.3 | 25 | 32.1 |
| Total | 150 | 100 | 72 | 100 | 78 | 100 |

### Once-Daily Brushing Is Predominant

The frequency of tooth brushing is shown in Figure 6 and Table 7. The majority (90.7%) brushed once daily, 5.3% brushed twice daily, and 4% brushed occasionally. More females (91%) than males (90.3%) brushed once daily, more females (4.2%) than males (4.2%) brushed twice daily, and more males (5.6%) than females (2.6%) brushed occasionally.

**Figure 6:**
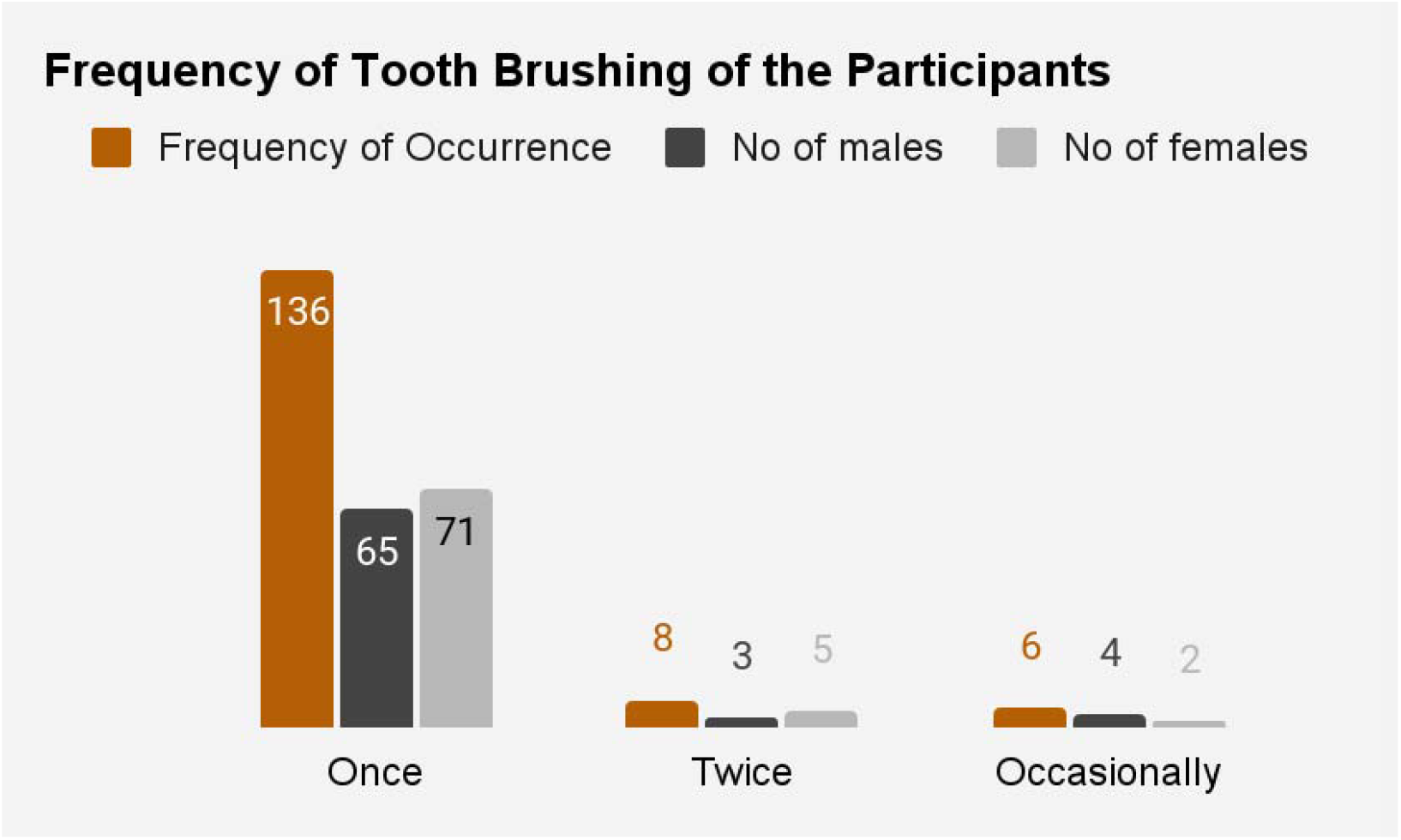
Frequency of Tooth Brushing. This figure illustrates the frequency of tooth brushing among the participants, indicating that 90.7% brush once daily, 5.3% brush twice daily, and 4% brush occasionally.

**Table 7:** Frequency of Tooth Brushing of the Respondents. This table illustrates how often participants brush their teeth, highlighting that once-daily brushing is the most common practice.

| Frequency of brushing | Frequency | % | No of males | % of males | No of females | % of females |
| --- | --- | --- | --- | --- | --- | --- |
| Once | 136 | 90.7 | 65 | 90.3 | 71 | 91 |
| Twice | 8 | 5.3 | 3 | 4.2 | 5 | 6.4 |
| Occasionally | 6 | 4 | 4 | 5.6 | 2 | 2.6 |
| Total | 150 | 100 | 72 | 100 | 78 | 100 |

### Dental Calculus Is the Most Common Oral Deposit

Oral deposits observed among participants are detailed in Figure 7 and Table 8. Dental plaque was observed in 16.7%, dental calculus in 47.3%, material alba in 20.7%, and dental stains in 14.7% of participants.

**Figure 7:**
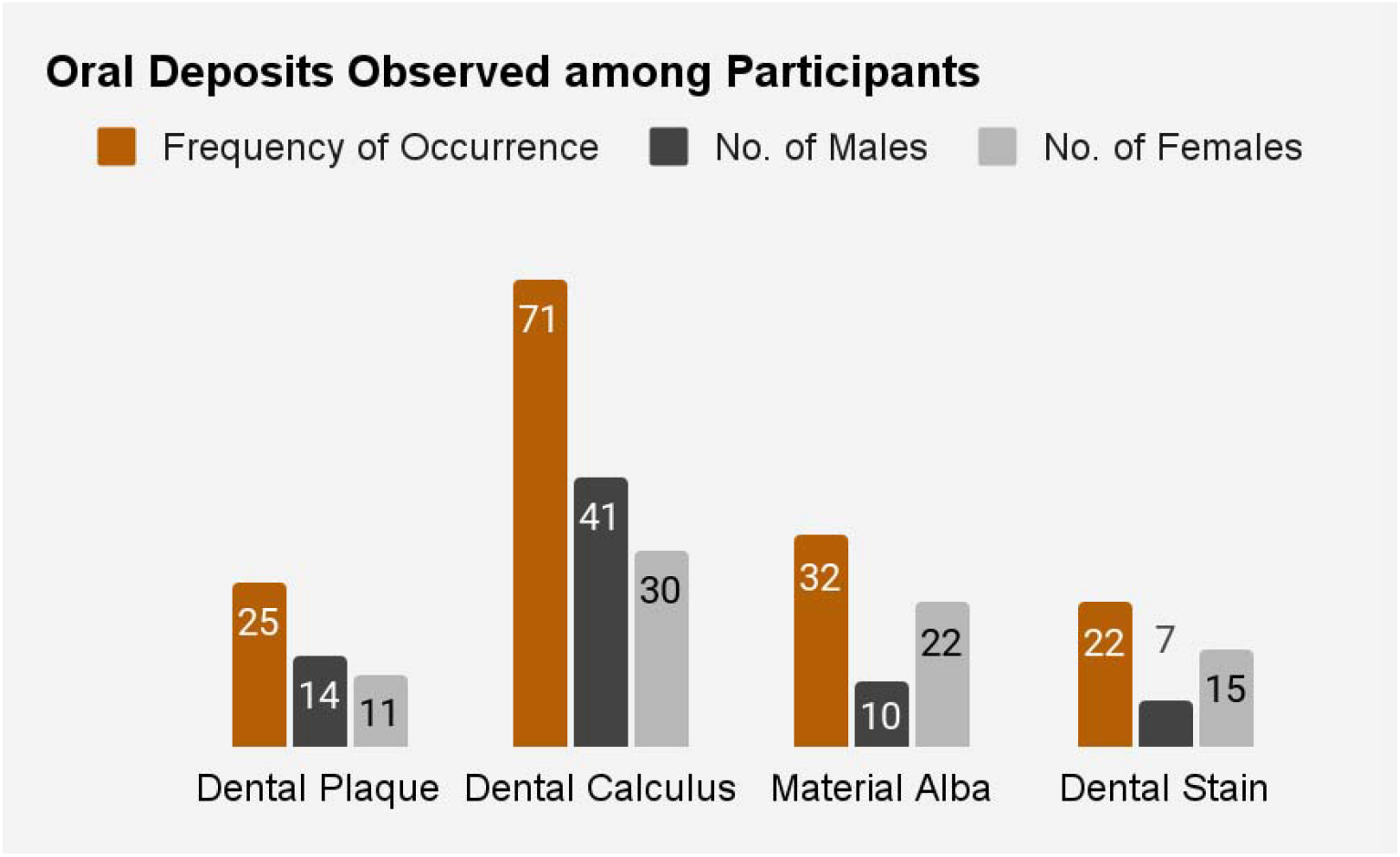
Oral Deposits Observed Among Participants. This figure presents the prevalence of various oral deposits, including dental plaque (16.7%), dental calculus (47.3%), material alba (20.7%), and dental stains (14.7%).

**Table 8:** Oral Deposits Observed. This table lists the types and frequencies of oral deposits found in participants’ mouths, including dental plaque, dental calculus, material alba, and dental stains.

| Oral Deposits | Frequency | % | No. of Males | % of Males | No. of Females | % of Females |
| --- | --- | --- | --- | --- | --- | --- |
| Dental Plaque | 25 | 16.7 | 14 | 19.4 | 11 | 14.1 |
| Dental Calculus | 71 | 47.3 | 41 | 56.9 | 30 | 38.5 |
| Material Alba | 32 | 20.7 | 10 | 13.9 | 22 | 28.2 |
| Dental Stain | 22 | 14.7 | 7 | 9.7 | 15 | 19.2 |
| Total | 150 | 100 | 72 | 100 | 78 | 100 |

### Dental Caries Are the Most Common Oral Disease

Common oral diseases observed among the participants are highlighted in Figure 8 and Table 9. Dental caries were found in 29.3% of participants, halitosis in 19.3%, gingivitis in 18%, and periodontitis in 10%. Only 23.3% of participants had no apparent oral disease but had oral deposits.

**Figure 8:**
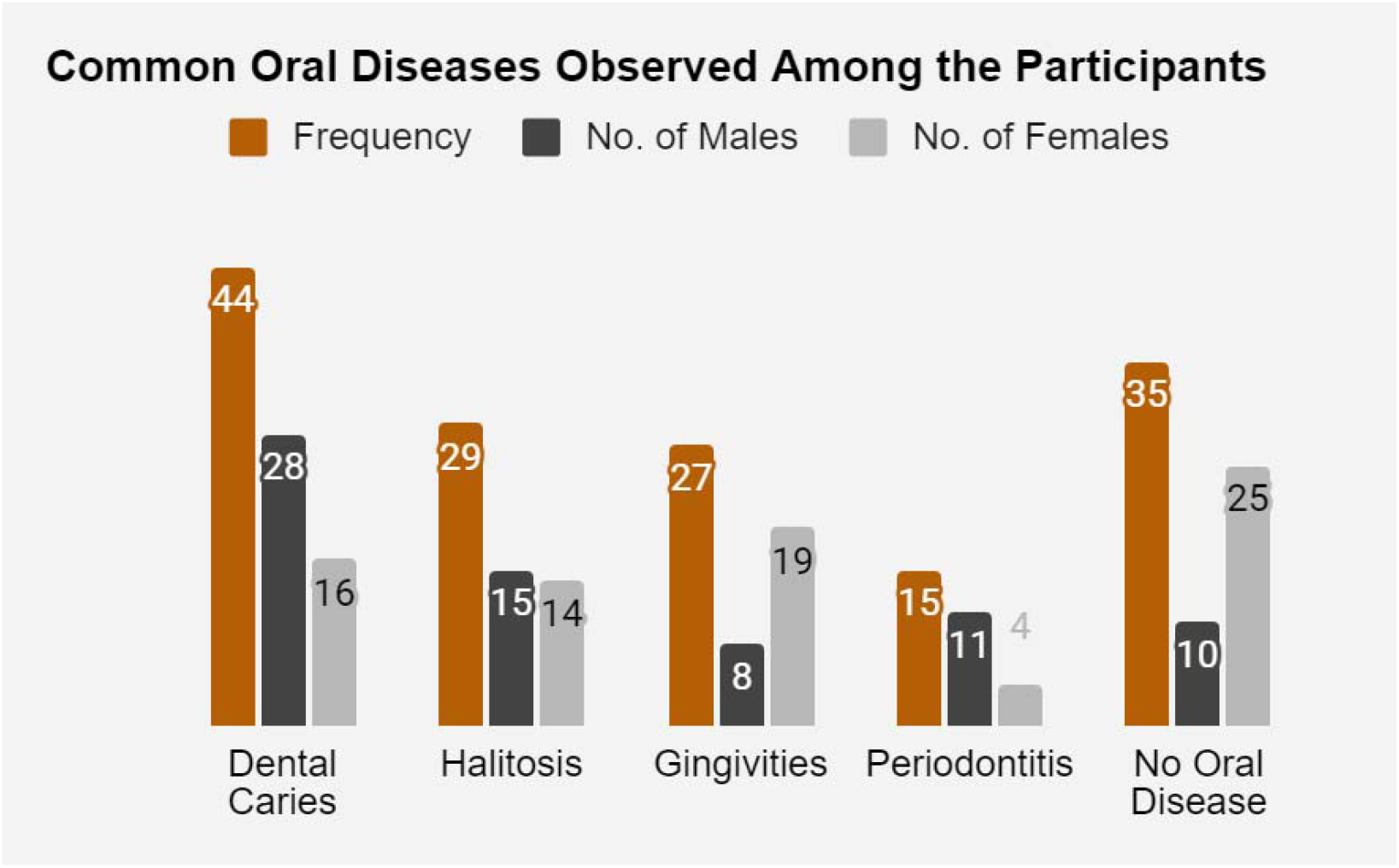
Common Oral Diseases Observed Among the Participants. This figure highlights the common oral diseases found in the participants, with dental caries (29.3%), halitosis (19.3%), gingivitis (18%), and periodontitis (10%) being the most prevalent.

**Table 9:** Common Oral Diseases Observed. This table provides data on the prevalence of various oral diseases among the participants, including dental caries, halitosis, gingivitis, and periodontitis.

| Oral Deposits | Frequency | % | No. of Males | % of Males | No. of Females | % of Females |
| --- | --- | --- | --- | --- | --- | --- |
| Dental Caries | 44 | 29.3 | 28 | 38.9 | 16 | 20.5 |
| Halitosis | 29 | 19.3 | 15 | 20.8 | 14 | 17.9 |
| Gingivitis | 27 | 18 | 8 | 5.3 | 19 | 24.4 |
| Periodontitis | 15 | 10 | 11 | 7.3 | 4 | 5.1 |
| No Oral Disease | 35 | 23.3 | 10 | 6.7 | 25 | 32.1 |
| Total | 150 | 100 | 72 | 100 | 78 | 100 |

## DISCUSSION

The cross-sectional study conducted in the Akpugo community of Enugu State, Nigeria, provided significant insights into the oral hygiene practices and their effectiveness in preventing oral diseases among the residents. Despite global improvements in oral health, this study reveals that oral diseases remain a significant public health concern in the Akpugo community [13].

The study found a high prevalence of oral diseases, with 29.3% of participants having dental caries, 19.3% suffering from halitosis, 18% diagnosed with gingivitis, and 10% with periodontitis. These findings indicate that a substantial portion of the population has unmet dental treatment needs. The prevalence of these conditions is likely due to limited access to dental care and an unequal distribution of healthcare resources, a common issue in underprivileged communities globally [14].

Most participants (90.7%) brushed their teeth once daily, predominantly in the morning, with only 5.3% brushing twice daily. This infrequent brushing practice contributes significantly to the high prevalence of oral diseases observed. Similar studies have reported poor oral hygiene practices due to infrequent tooth cleaning, particularly after consuming sugary snacks, which can lead to dental caries [15]. The slight female dominance in twice-daily brushing aligns with previous research indicating that females generally have better oral hygiene habits. This can be attributed to higher health awareness and more rigorous personal care routines among women [16].

The preference for toothbrushes and toothpaste (58%) over traditional methods such as chewing sticks (26.67%) reflects a shift towards modern oral hygiene practices. However, the continued use of chewing sticks, particularly among older age groups, suggests that traditional practices still play a role in the community’s oral hygiene regimen. The use of modern tools over traditional methods may be influenced by economic conditions, governance, and increased access to information [17].

The study’s findings that 56% of participants brushed their teeth only in the morning highlight a significant gap in effective oral hygiene practices. This practice, coupled with the low rate of twice-daily brushing, underscores the need for better oral health education and the promotion of more frequent brushing habits to reduce the incidence of oral diseases [18].

The observation of dental calculus in 47.3% of participants and dental plaque in 16.7% indicates poor oral hygiene practices. The presence of these deposits correlates strongly with the high prevalence of dental caries, gingivitis, and periodontitis observed in the study. Effective oral hygiene measures, such as regular brushing and professional dental cleanings, are essential to reduce these deposits and improve overall oral health [19].

The disparity in oral hygiene practices between the Akpugo community and other regions, such as Scotland, where a higher percentage of the population practices twice-daily brushing, highlights significant differences in healthcare infrastructure and education. This disparity underscores the need for tailored interventions that address the specific needs and practices of rural communities like Akpugo [20].

The study’s limitations include a small sample size of 150 participants, which may not fully represent the diversity of the Akpugo community. The reliance on self-reported data introduces potential biases, and variations in examiners’ interpretations could affect the accuracy of oral health assessments. Additionally, cultural and socioeconomic factors influencing oral health practices were not extensively explored, limiting the depth of understanding [21].

To improve oral health outcomes in the Akpugo community, targeted interventions are necessary. Comprehensive oral health education programs should be developed to promote proper tooth brushing techniques and raise awareness about the importance of regular dental check-ups. Collaboration with local healthcare authorities and NGOs is essential to establishing dental centers or clinics within the community, ensuring access to affordable oral health services. Empowering community members through oral health campaigns and seminars can foster sustainable improvements in oral hygiene practices. Longitudinal studies are recommended to monitor changes in oral health behaviors over time and evaluate the impact of interventions. Collaborative research involving multidisciplinary teams can enrich future endeavors, leading to more comprehensive strategies for addressing oral health disparities and promoting oral health equity in the Akpugo community [22].

These initiatives are essential for reducing the prevalence of oral diseases and enhancing the overall health and well-being of the Akpugo community. The study highlights the importance of adopting effective oral hygiene practices and underscores the necessity for continuous education and intervention to promote optimal oral health in rural communities. Through comprehensive assessment and targeted educational initiatives, the research aims to empower the community with the knowledge and tools required to maintain good oral health and improve their quality of life [23].

## Data Availability

All data produced in the present work are contained in the manuscript

